# The localized rise of a B.1.526 SARS-CoV-2 variant containing an E484K mutation in New York State

**DOI:** 10.1101/2021.02.26.21251868

**Authors:** Erica Lasek-Nesselquist, Pascal Lapierre, Erasmus Schneider, Kirsten St. George, Janice Pata

## Abstract

The E484K mutation in the spike protein of SARS CoV-2 contributes to immune escape from monoclonal antibodies as well as neutralizing antibodies in COVID-19 convalescent plasma. It appears in two variants of concern – B.1.351 and P.1 - but has evolved multiple times in different SARS-CoV-2 lineages, suggesting an adaptive advantage. Here we report on the emergence of a 484K variant in the B.1.526 lineage that has recently become prevalent in New York State, particularly in the New York City metropolitan area. In addition to the E484K mutation, these variants also harbor a D235G substitution in spike that might help to reduce the efficacy of neutralizing antibodies.

## Introduction

In recent months, several SARS-CoV-2 variants of concern have appeared internationally – notably B.1.1.7 (501Y.V1), B.1.351 (501Y.V2), and P.1 (501Y.V3). Variant B.1.1.7 was estimated to have emerged in late September 2020 and rapidly became the dominant lineage in England, where it was first identified (Rambaut et al. 2020; Volz et al. 2021). Similarly, B.1.351 has quickly become the dominant lineage in many parts of South Africa, where it was initially detected in October 2020 (Tegally et al. 2020). Variant P.1 was first identified in four travelers from Brazil and is associated with a case of reinfection (Faria et al. 2021; Naveca et al. 2021). All three variants or lineages of concern harbor a suite of mutations in the spike protein, which is required for host cell entry by binding to the angiotensin-converting enzyme-2 (ACE-2) receptor. In particular, B.1.1.7, B.1.351, and P.1 share an N501Y mutation that increases ACE2 receptor binding affinity (Chan et al. 2020; Starr et al. 2020) and cell infectivity in animal models (Gu et al. 2020) and likely contributes to the increased transmission observed for B.1.1.7 and B.1.351 (Tegally et al. 2020; Davies et al. 2021; Volz et al. 2021). The mutations K417N and E484K found in B.1351 and P.1 dramatically reduce the neutralization activity of monoclonal antibodies and convalescent plasma/sera (Weisblum et al. 2020; Cele et al. 2021; Liu et al. 2021; Wibmer et al. 2021; Greaney et al. 2021), making them of considerable concern for vaccine efficacy.

The largely uncontrolled spread of SARS-CoV-2, particularly in the United States (US), increases the risk for the rise of additional variants that might have selective advantages over ancestral types. For example, B.1.429 – defined by an L452R mutation in spike – was responsible for 24% of all COVID-19 cases in Southern California by November after its first detection in July (Zhang et al. 2021). L452R might increase infectivity by stabilizing the spike-ACE2 receptor interaction and has evolved independently in multiple lineages (such as B.1.427; Tchesnokova et al. 2021). While surveillance efforts have increased in the US, the country has not yet met the recommended sequencing of 5% of all COVID-19 cases in order to detect variants before they reach high prevalence in the population (Vavrek et al. 2021).

Here we report on the emergence of a 484K variant within the B.1.526 lineage that has increased in the circulating virus population in New York state by almost 26-fold in a little over a month. Since its first detection in New York on 12-16-2021, E484K is now one of the most frequently identified spike mutations in SARS-CoV-2 genomes in the state. B.1.526 typically has five other amino acid substitutions in the spike protein - L5F, T95I, D253G, D614G, and A701V. D253 falls in the “N5” or “supersite” loop of the N-terminal domain (NTD) of spike (Cerutti et al. 2021; McCallum et al. 2021) and was identified as one of the two key residues (the other being R246) that interact with most NTD-reacting neutralizing antibodies (Wibmer et al. 2021). Thus, D253G may be akin to the R246I mutation seen in B.1.351 by abolishing an important neutralization epitope and contributing to antibody neutralization escape.

Additionally, an S477N mutation has arisen independently twice within the B.1.526 lineage. S477N is associated with increased ACE2 receptor binding affinity (Zahradník et al. 2021) and resistance to neutralization by monoclonal antibodies (Liu et al. 2021) but other lineages containing this mutation (such as 20A.EU2) are not dramatically increasing in New York. The combination of E484K or S477N with a D253G mutation that might confer immune escape and the increased number of COVID-19 cases associated with these variants warrants further monitoring.

## Results

Mutations in more than 10,000 SARS-CoV-2 genomes sampled from New York were cataloged by NextClade (https://clades.nextstrain.org/). Since December 2020, we observed a dramatic increase in the number of genomes that contained spike E484K mutations (Figure 1), which predominantly segregated with five other spike mutations that characterize the B.1.526 lineage - L5F, T95I, D253G, D614G, and A701V.

**Figure 1.**
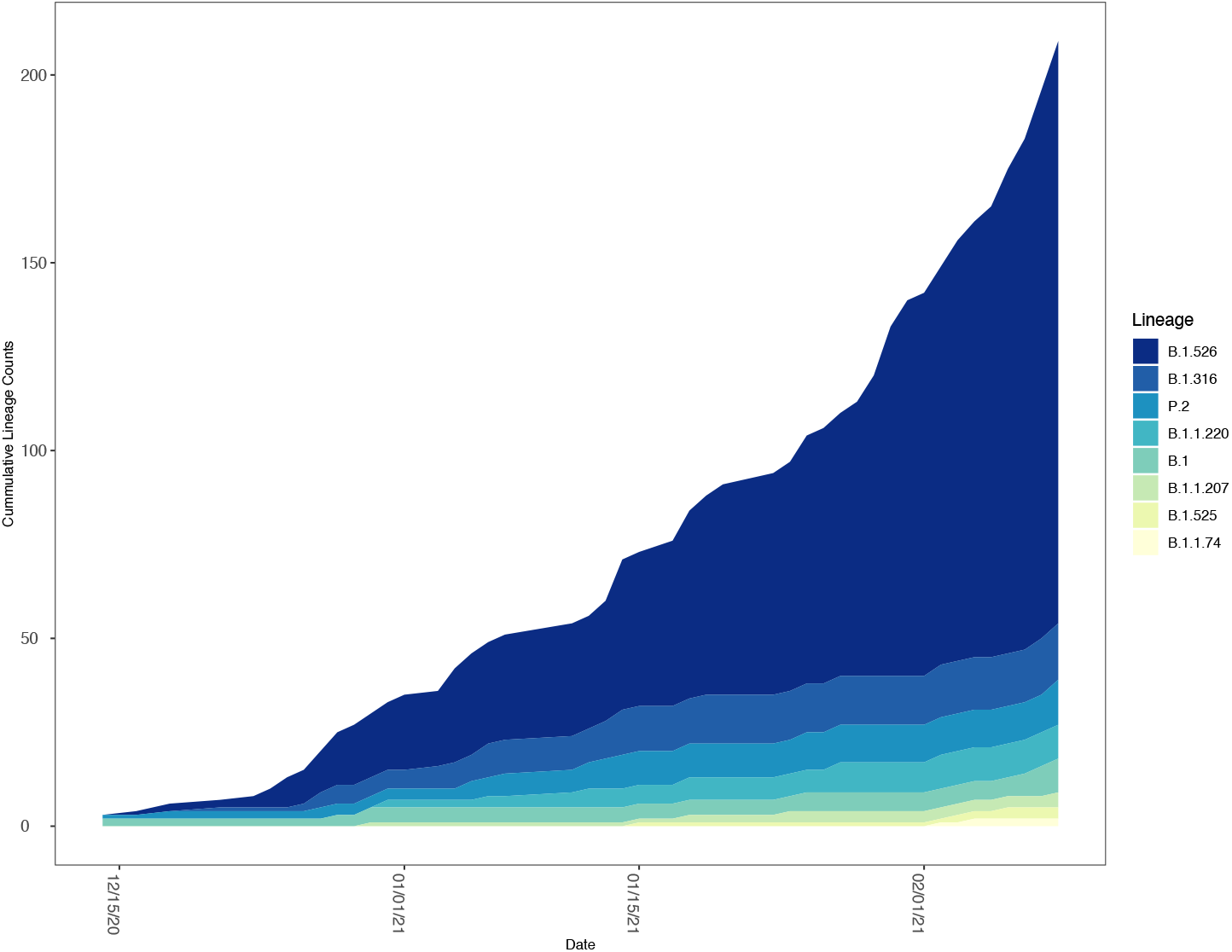
A) Relative frequency of SARS-CoV-2 lineages with E484K mutations in New York State. The number of SARS CoV-2 genomes assigned to lineages containing E484K as a function of time. The lineages included contained at least one genome with an E484K mutation in the New York State dataset.

**Figure 1.**
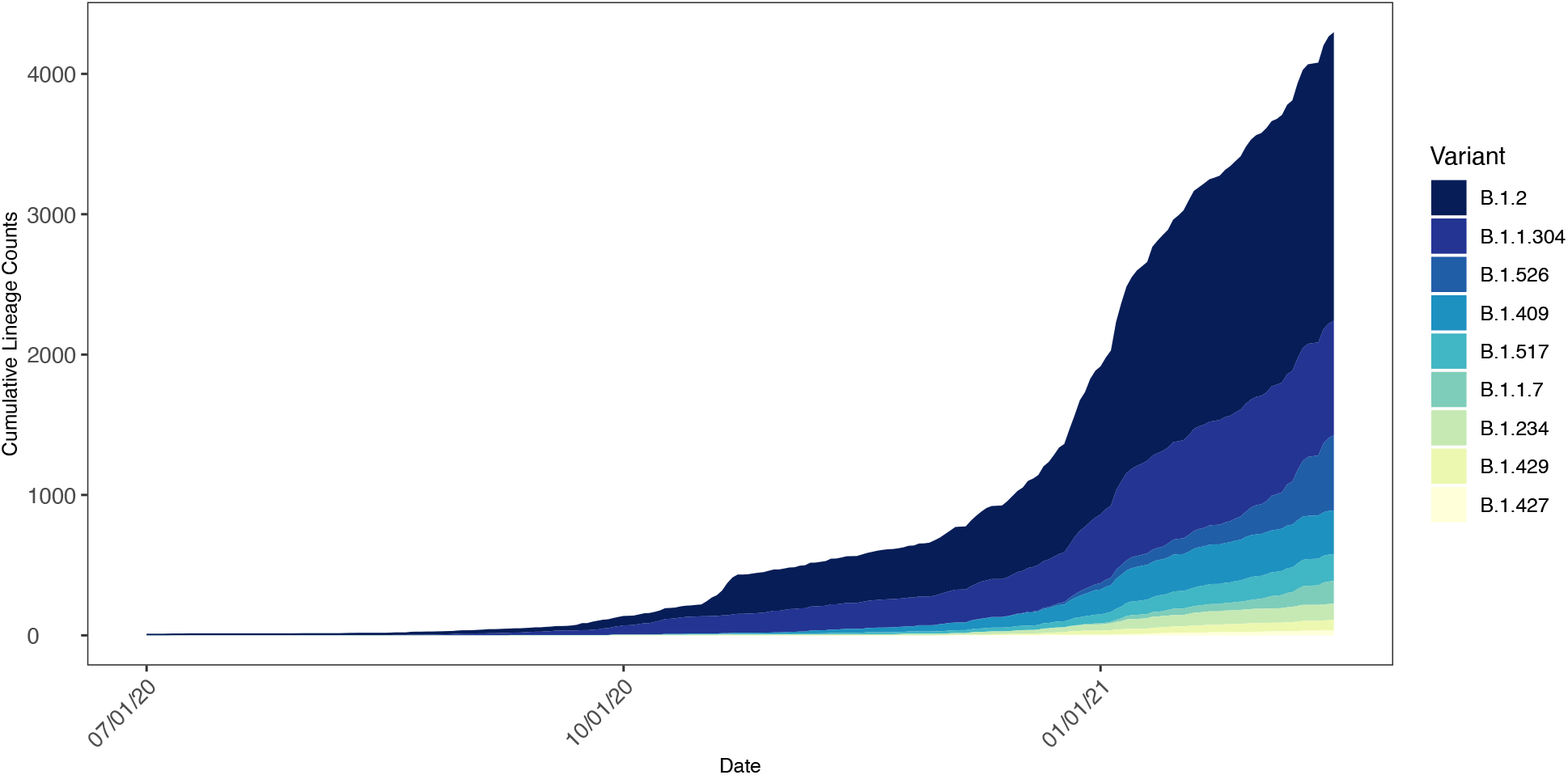
B) Changing frequency of B.1.526 and other lineages in New York State. The number of genomes assigned to select SARS-CoV-2 lineages circulating in New York State as a function of time. Only lineages with 10 or more genomes from samples collected within the past 14 days were included.

As of 02-22-2021, there were 579 B.1.526 genomes deposited to GISAID, 535 of which (92%) derived from New York State, almost exclusively (99%) from the larger New York City metropolitan area (the five boroughs of NYC, and the Long Island and Mid-Hudson regions; Figure 2). The remaining 8% were predominantly from the Northeastern United States.

**Figure 2.**
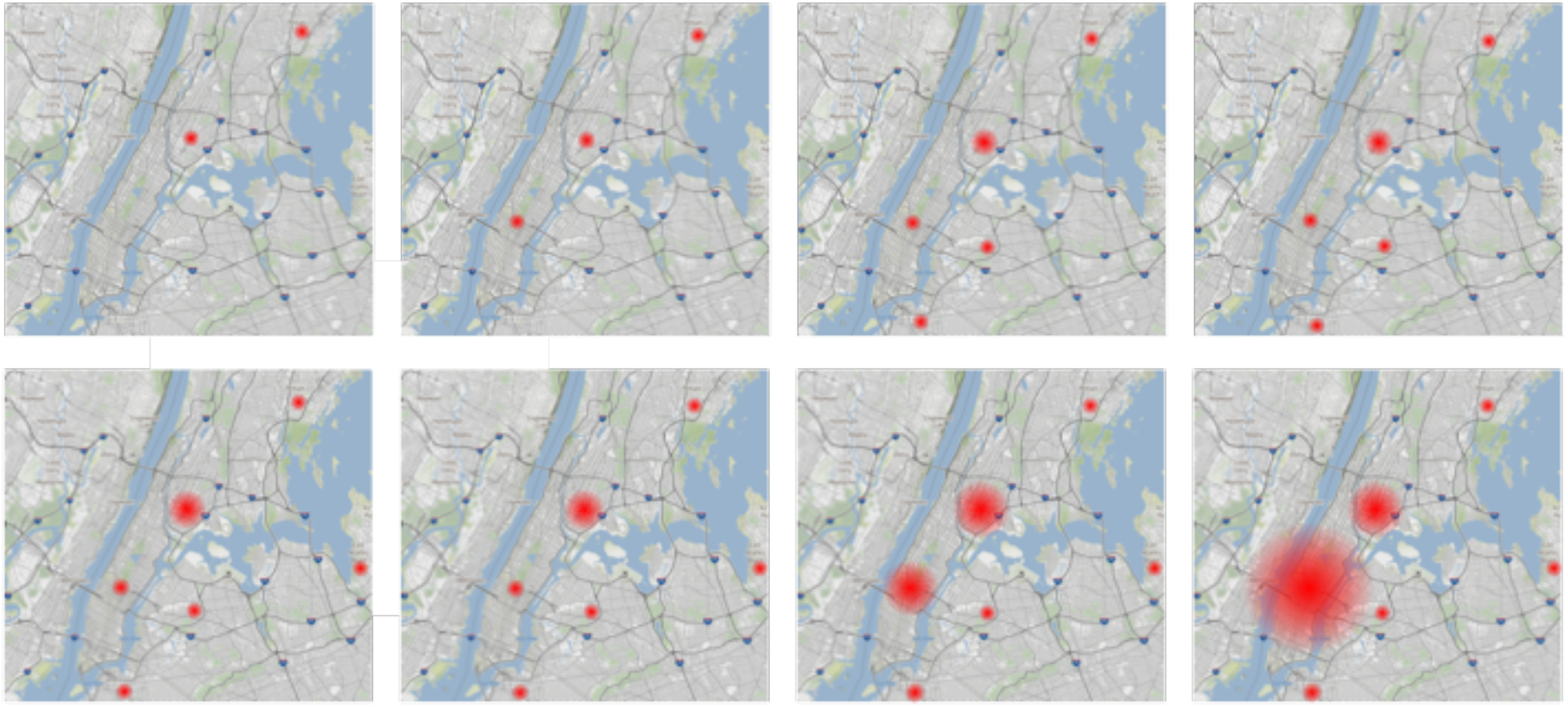
Increase in Frequency of the B.1.526 E484K variant in the NYC metropolitan area. Starting from the top left, each panel encompasses a seven-day period from 2020-12-16 to 2021-02-03 showing the increase of B.1.526 484K variants (red dots). The red dots are proportional to the total number of variants found in an area.

The first SARS-CoV-2 genome assigned to the B.1.526 lineage and the subsequent acquisition of the E484K mutation were identified in New York on 2020-11-23 and 2020-12-16, respectively. Prior to the first appearance of the E484K mutation was the detection of a B.1.526 SARS CoV-2 genome with an S477N mutation (collected on 2020-12-08). Thus, both 484K and 477N variants appear to have arisen independently on the same mutational background.

To estimate the timing of introductions and geographical origins of 484K and 477N variants, we downloaded all B.1.526 genomes as well as contextual sequences from GISAID and performed a phylogeographic analysis using the ‘ncov’ pipeline from Nextstrain (Hadfield et al. 2018; Figure 3). Phylogeographic analyses suggested that the 484K variant emerged in New York by mid-November (Figure 3; 100% CI, 2020-10-21 to 2020-12-11). Interestingly, 477N appears to have evolved twice independently from the parental B.1.526 (E484, S477) lineage, with the largest cluster emerging in New York by early December (Figure 3; 100% CI, 2020-11-26 to 2020-12-24).

**Figure 3.**
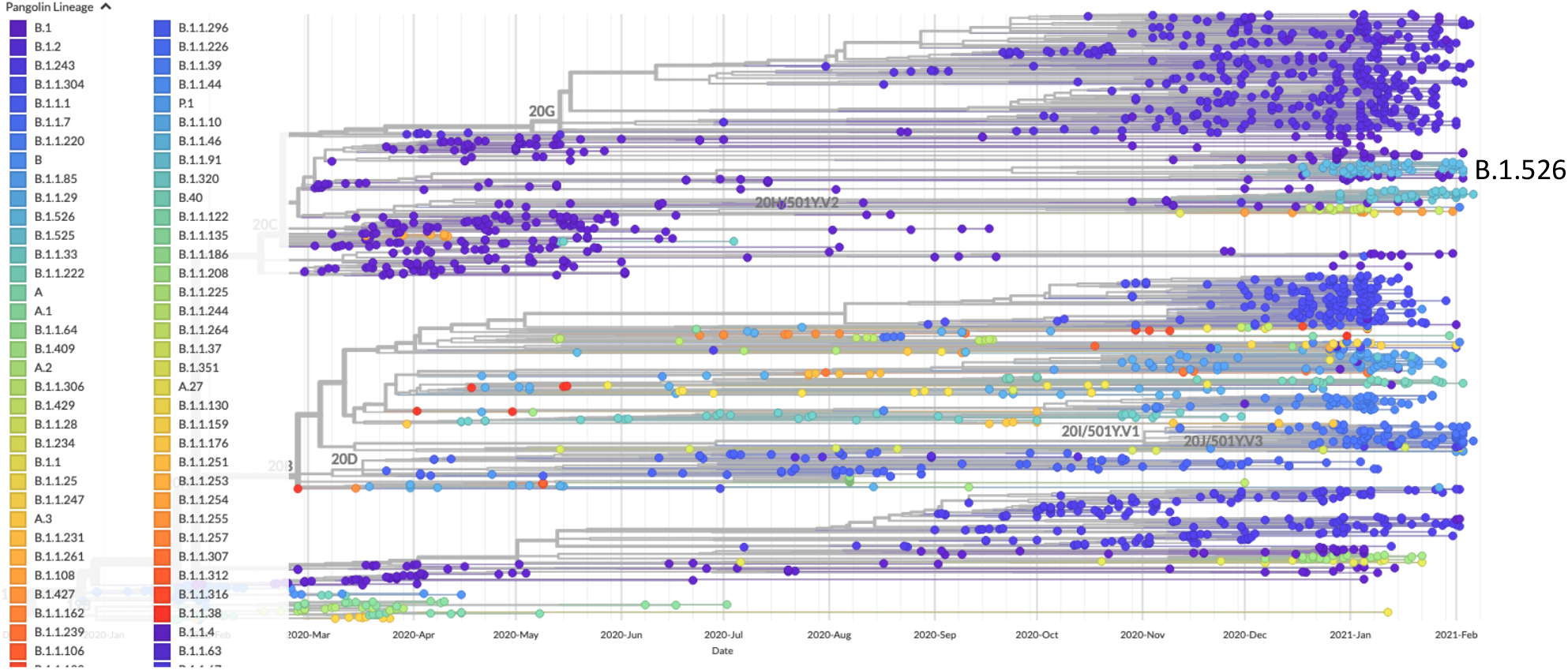
A) Time-resolved phylogeny of SARS-CoV-2. A global phylogeny of SARS-CoV-2 generated in Nextstrain and visualized in Auspice to show the phylogenetic placement of the B.1.526 lineage.

**Figure 3.**
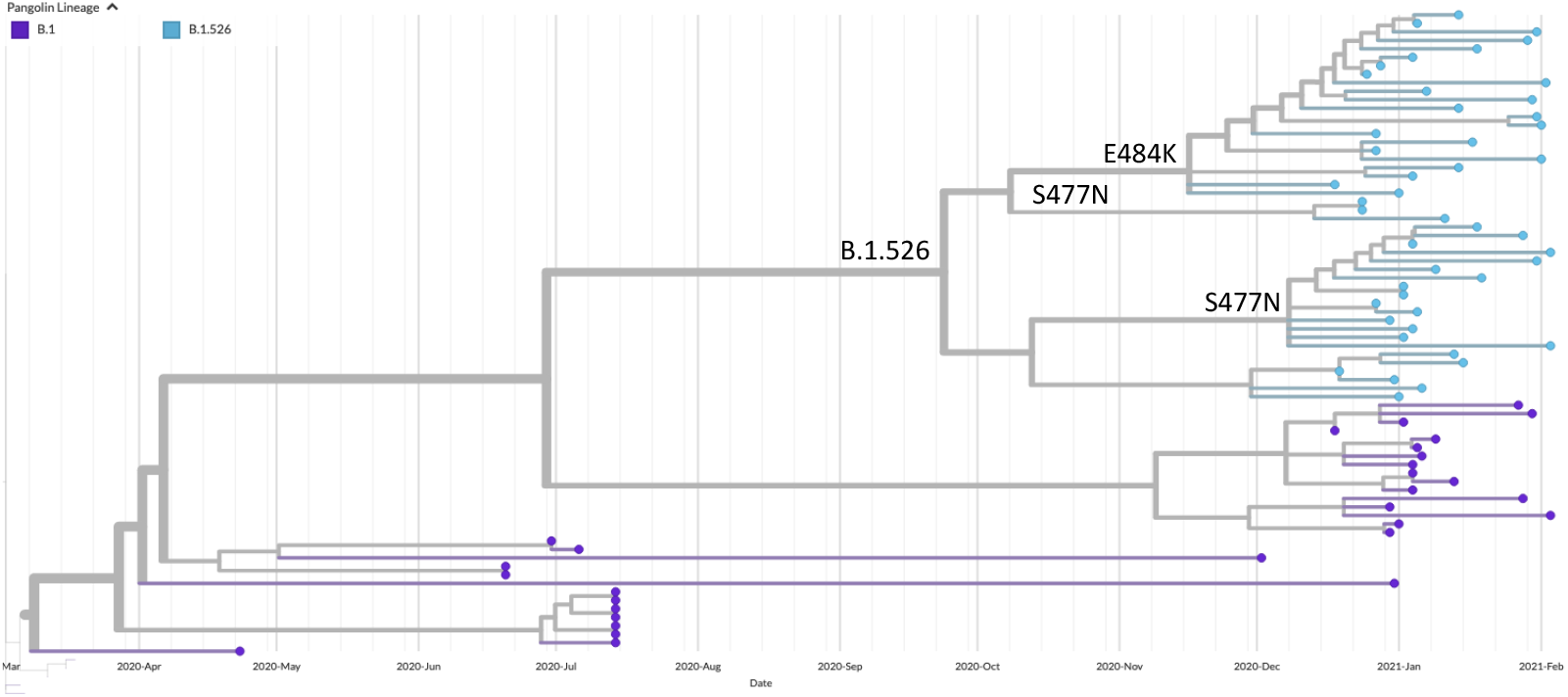
B) Subtree of B.1.526 lineage. A subtree containing the B.1.526 lineage with the clades containing E484K or S477N mutations identified to highlight their recent emergence.

The 484K and 477N variants now co-circulate with the parental B.1.526 lineage. However, both spike variants currently comprise the majority of B.1.526 cases (434/535 or 81%; Figure 4).

**Figure 4.**
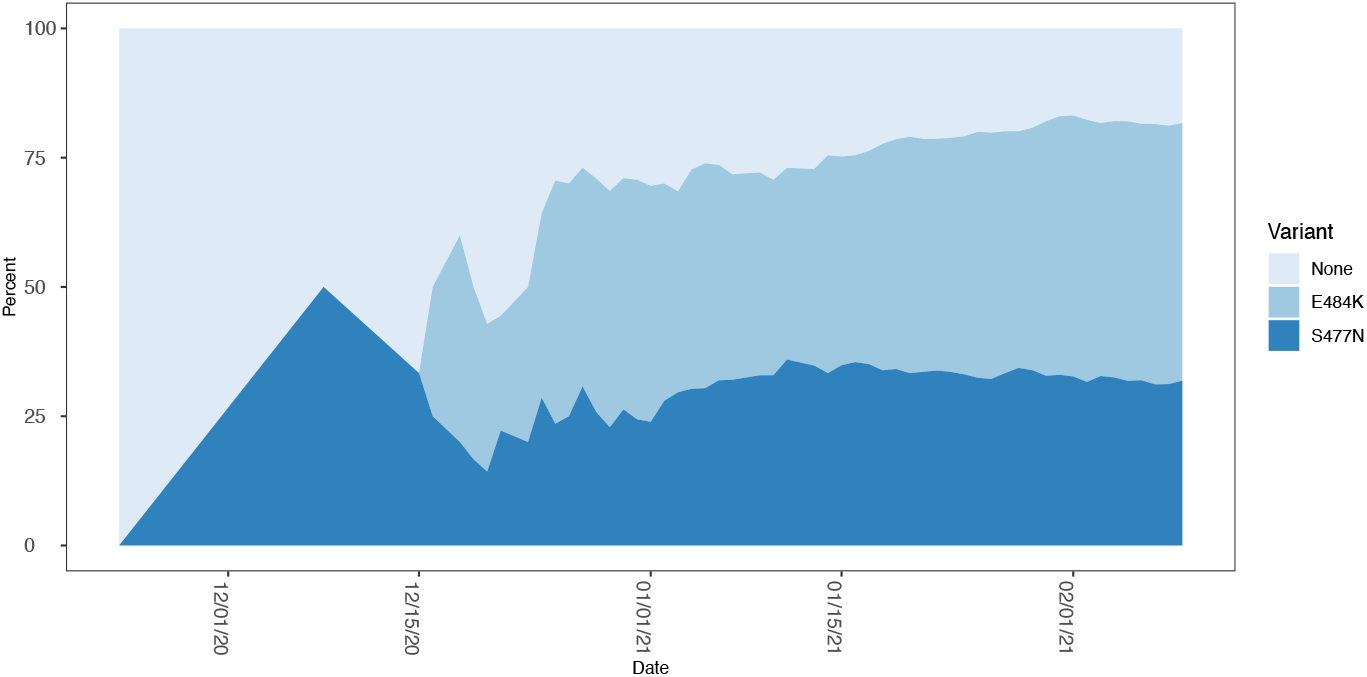
Relative proportion of B.1.526 parental and variant lineages in New York. Stacked area map of the proportion of B.1.526 genomes that harbor an E484K, S447N, or neither (None) mutation as a function of time.

We also downloaded all genomes with an E484K mutation to compare the frequency of B.1.526 484K variants to other lineages with this amino acid change. As of 2021-02-22, the B.1.526 lineage was the third most abundant lineage with an E484K mutation, surpassed only by B.1.351 and P.2 and exceeding B.1.525 and A.23.1 (Table 1), which are listed among the variants of concern on the Pango Lineages website (https://cov-lineages.org/index.html).

**Table 1.** The number of genomes with E484K mutations per lineage. Other is the cumulative count of E484K mutations for 70 lineages.

| Lineage | Count |
| --- | --- |
| B.1.351 | 1802 |
| P.2 | 524 |
| **B.1.526** | **304** |
| Other | 301 |
| B.1.525 | 207 |
| P.1 | 173 |
| R.1 | 118 |
| B.1.1.207 | 111 |
| B.1 | 106 |
| A.23.1 | 63 |
| B.1.1.7 | 47 |

E484K showed the largest increase in frequency from 2021-01-13 to 2021-02-22 in comparison to other mutations of interest circulating in New York, surpassing N501Y (associated with B.1.1.7) and L452R (associated with B.1.429/B.1.427; Table 2). In fact, both E484k and S477N mutations showed the greatest fold change out of all mutations of biological significance considered, either in total or associated with B.1.526 (Table 2).

**Table 2.** Increase in mutations of interest in New York. The percentage of SARS-CoV-2 genomes from New York State with a mutation of interest as of 2021-01-13 and 2021-02-22 (number of genomes in parentheses) and the fold change between the two sampling dates. Collection date, the collection date associated with the first appearance of each mutation in the New York State. E484K and S477N were first identified around the same time or after other mutations of interest, suggesting that differences in starting dates do not strongly influence the trends observed. E484K (B.1.526) and S477N (B.1.526) are only the E484K and S477N mutations associated with the B.1.526 lineage as opposed to any lineage (E484K and S477N). The first occurrence of N501Y is in April 2020 but is not associated with B.1.1.7 and was not included.

| Mutation | 2021-01-13 | 2021-02-22 | Fold increase | Collection date |
| --- | --- | --- | --- | --- |
| E484K (B.1.526) | 0.43 (35) | 11 (243) | 26 | 2020-12-16 |
| E484K | 0.65 (53) | 13 (276) | 20 | 2020-12-16 |
| S477N | 0.34 (28) | 5.5 (118) | 16 | 2020-12-08 |
| S477N (B.1.526) | 0.37 (30) | 5.8 (126) | 16 | 2020-12-08 |
| N501Y | 0.41 (33) | 5.9 (127) | 14 | 2020-12-24 |
| L452R | 1.0 (83) | 8.3 (179) | 8.3 | 2020-10-29 |
| N501T | 1.5 (125) | 3.3 (71) | 2.1 | 2020-09-09 |
| P681H | 9.0 (734) | 19 (405) | 2.1 | 2020-01-30 |
| Total number sequences | 8140 | 2162 |  |  |

## Discussion

We note the emergence of new variants in the B.1.526 lineage, which now harbor E484K or S477N mutations in addition to the already present D253G change. Collectively, these variants represent 81% of B.1.526 SARS-CoV-2 genomes in New York State, helping to make it the fifth most abundant lineage since its detection in mid-December. In fact, the number of B.1.526 genomes is only surpassed by B.1 and B.1.2, which have been circulating in the population since early in the pandemic, and B.1.243 and B.1.1.304 – also in circulation months earlier than B.1.526. The E484K mutation has been shown to reduce the efficacy of neutralizing antibodies and it is hypothesized that 253G could do the same as it is located in a short, disordered region of the “supersite” loop of the NTD, near the interface with the neutralizing antibody 4A4 (Figure 5, right panel; Chi et al. 2020). Additionally, a mutation at E484 could alter binding affinity and potentially contribute to the dramatic rate of increase of B.1.526 484K variants in New York as it is located in the RBD, at the interface with the ACE2 receptor (Figure 5, left panel; Yan et al. 2020). Other mutations present in B.1.526 likely do not explain the increase in transmission. T95 is a buried residue in the NTD, and thus T95I would not seem to affect antigenicity. It is also not proximal to the receptor binding domain (RBD) in a neighboring subunit in the trimer, so the relatively conservative T to I substitution probably does not influence the RBD closed-to-open transition (Paul Masters, personal communication). Neutralization studies of A701V have looked at this mutation only in conjunction with multiple other mutations in B.1.351 (Wibmer et al. 2021; Cele et al. 2021) so its contribution to phenotype has not been assessed.

**Figure 5.**
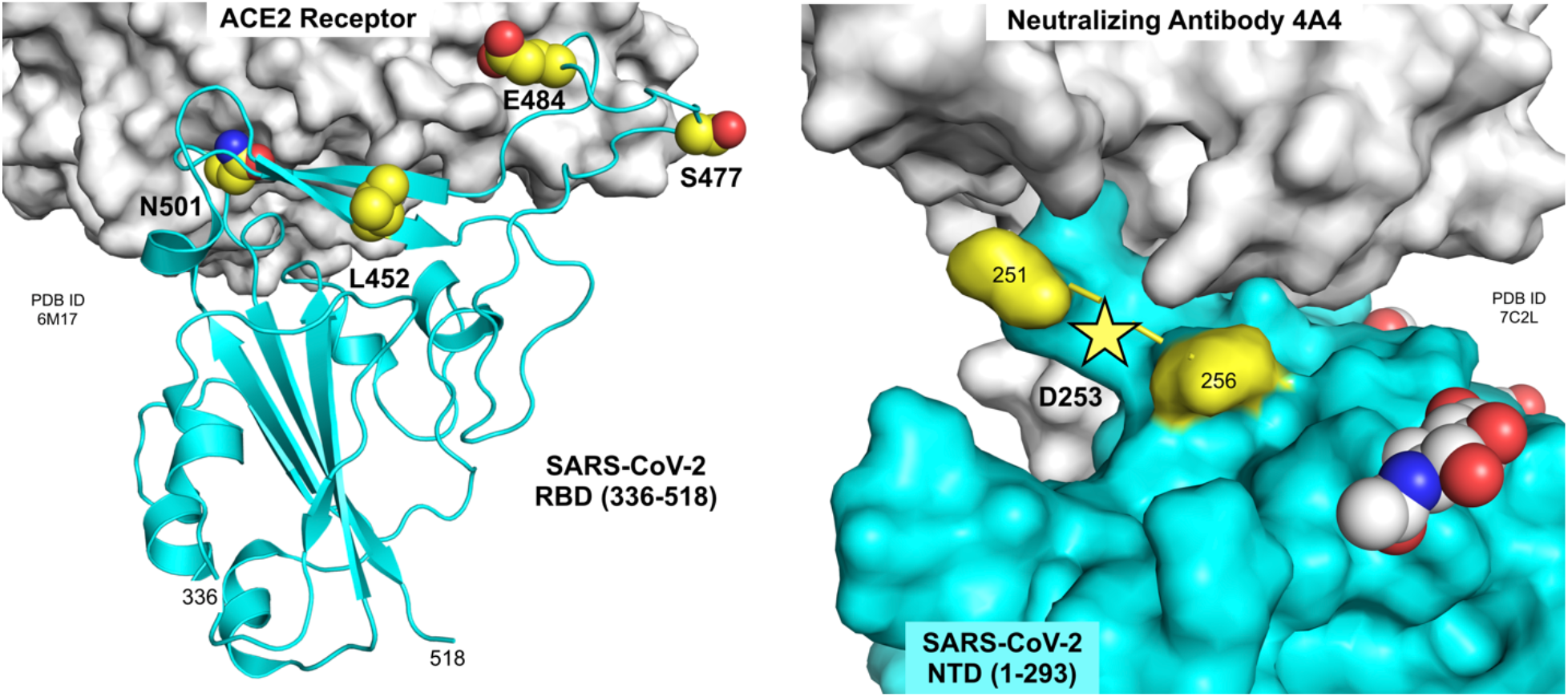
Residues mutated in B.1.526 spike protein. Residues 452, 477, 484 and 501 are located at or near the interface of the RBD with the ACE2 receptor (left panel; PDB ID 6M17, Yan et al., 2020). Residue 253 is located in a small disordered loop in the NTD near the interface with neutralizing antibody 4A4 (right panel; PDB ID 7C2L, Chi et al., 2020). Figure prepared using the PyMOL Molecular Graphics System v2.3.5 (Schrödinger, LLC).

There are also caveats to consider. The increase in frequency of 484K and 477N variants might be due to stochastic processes unrelated to phenotype. While we would expect an initial localized spike in the number of variants that confer a selective advantage, we have not observed a substantial spread of either B.1.526 484K or 477N from the NYC metropolitan area to other regions of the state since their first detection. However, the combination of two mutations that potentially provide immune escape in a SARS-CoV-2 lineage that has rapidly risen in frequency warrants further monitoring.

## Methodology

### RNA extraction

Respiratory swabs in viral transport medium (VTM, UTM or MTM) were received at the Virology Laboratory, Wadsworth Center, for SARS-CoV-2 diagnostic testing. Total nucleic acid was extracted using either easyMAG or eMAG (bioMerieux, Durham, NC), MagNA Pure 96 and Viral NA Large Volume Kit (Roche, Indianapolis, IN), or EZ1 with the DSP Virus Kit (QIAGEN, Hilden, Germany) following manufacturers’ recommendations. Extracted nucleic acid was tested for SARS-CoV-2 RNA using the 2019-Novel Coronavirus Real-Time RT-PCR Diagnostic Panel (Centers for Disease Control and Prevention) according to the Instructions for Use on an ABI 7500Dx.

### Illumina library preparation and sequencing

Whole genome amplicon sequencing of SARS-CoV-2 was performed using a modified ARTIC protocol (https://artic.network/ncov-2019). Briefly, cDNA was synthesized by combining 5 µl of RNA with SuperScript™ IV reverse transcriptase (Invitrogen, Carlsbad, CA, USA), random hexamers, dNTPs, and RNAse inhibitor, followed by incubation at 25 °Cfor 5 minutes, 42 °Cfor 50 minutes and 70 °Cfor 10 minutes on a SimpliAmp thermal cycler (Thermo Fisher Scientific, Waltham, MA, USA). Two separate amplicon pools were generated by multiplex PCR with two separate premixed ARTIC V3 primer pools (Integrated DNA Technologies, Coralville, IA, USA). Additional primers to supplement those that showed poor amplification efficiency (https://github.com/artic-network/artic-ncov2019/tree/master/primer_schemes/nCoV-2019) were added separately to the pooled stocks. PCR conditions were 98 °Cfor 30 seconds, followed by 24 cycles of 98 °Cfor 15 seconds and 63 °Cfor 5 minutes with a final 65 °Cextension for 5 minutes. Amplicons from pool 1 and pool 2 reactions were combined and purified by Ampure XP beads (Beckman Coulter, Brea, CA, USA) with a 1X bead to sample ratio and eluted in 10 mM Tris-HCl (pH 8.0). The amplicons were quantified using Quant-IT™ dsDNA Assay Kit on an ARVO™ X3 Multimode Plate Reader (Perkin Elmer, Waltham, MA, USA). Illumina sequencing libraries were generated using the Nextera DNA Flex Library Prep Kit with Illumina Index Adaptors and sequenced on a MiSeq or NextSeq instrument (Illumina, San Diego, CA, USA).

### Bioinformatics processing

Illumina libraries were processed with ARTIC nextflow pipeline (https://github.com/connor-lab/ncov2019-artic-nf/tree/illumina, last updated April 2020). Briefly, reads were trimmed with TrimGalore (https://github.com/FelixKrueger/TrimGalore) and aligned to the reference assembly MN908947.3 (strain Wuhan-Hu-1) by BWA (Li & Durbin 2009). Primers were trimmed and consensus sequences were generated with iVar (Grubaugh et al. 2018). Positions were required to be covered by a minimum depth of 50 reads and variants were required to be present at a frequency ≥ 0.75.

### Phylogeographic analysis

We constructed a time-resolved phylogeny of SARS-CoV-2 genomes in Nexstrain v2.0.0 (Hadfield et al. 2018) using representatives of the B.1.526 lineage sequenced by the Wadsworth Center and contextual sequences sampled from New York and the rest of the world (see Acknowledgements table for genomes used and contributing laboratories). In summary, a total of 2155 genomes >27 Kb in length were aligned in MAFFT v7.475 (Katoh & Standley 2013), a maximum likelihood phylogeny was generated with IQ-TREE v2.0.3 (Nguyen et al. 2015) under a GTR+G4 nucleotide substitution model, and divergence times as well as ancestral traits were inferred with TreeTime (Sagulenko et al. 2018). Trees were visualized in Auspice (Hadfield et al. 2018).

### IRB Approval

This work was approved by the New York State Department of Health Institutional Review Board, under study numbers 02-054 and 07-022.

## Supporting information

Acknowledgement table

## Data Availability

Genome sequences and metadata are available at GISAID.org

## Acknowledgements

Next generation sequencing was performed by the Advanced Genomic Technologies Core of the Wadsworth Center. Initial funding for sequencing was generously provided by the New York Community Trust. This publication was also supported by Cooperative Agreement number NU50CK000516, funded by the Centers for Disease Control and Prevention. Its contents are solely the responsibility of the authors and do not necessarily represent the official views of the Centers for Disease Control and Prevention or the Department of Health. We graciously thank all originating and submitting laboratories for their SARS-CoV-2 sequence contributions to the GISAID database, the Wadsworth Center’s Bioinformatics Core, and Paul Masters for his invaluable expertise.

